# Reduced central alpha power at rest is associated with risk of alcohol-related blackout and frequency of non-REM parasomnia episodes

**DOI:** 10.1101/2025.09.25.25336484

**Authors:** Grace M. Elliott, Madeline M. Robertson, Celine E. Locklear, Donita L. Robinson, Margaret A. Sheridan, Charlotte A. Boettiger

## Abstract

People who report experiencing alcohol-related blackouts (ARBs) are at increased risk of alcohol-related injury and even death. Blackout susceptibility is heritable and blackouts are not experienced by all who engage in hazardous drinking. Blackout is defined by anterograde amnesia, but a person in the blackout state also maintains consciousness and motor control at high levels of intoxication, which is behaviorally similar to episodes seen in individuals with a history of sleepwalking or related parasomnias. Spectral analysis of resting-state electroencephalograms (EEG) can provide insight into individual differences in baseline neurophysiology which may predict blackout susceptibility in otherwise healthy individuals. The current study investigated potential neurophysiological phenotypes present in the resting-state EEG spectra of individuals with a history of blackout, sleepwalking, or related parasomnias. In Experiment 1, adult females with a history of alcohol-related blackout had reduced resting-state alpha peak power over the primary motor cortex compared to those with no such history, while aperiodic slope over the right primary motor cortex was negatively correlated with lifetime blackout score in males. In Experiment 2, increased frequency of parasomnia episodes was associated with reduced resting-state alpha peak power across males and females. Together, these findings provide the first support for the existence of common neurophysiological phenotypes between specific parasomnias and alcohol-related blackout.

**New & Noteworthy:** Research on blackout often focuses on hippocampal suppression by alcohol because anterograde amnesia is a salient and definitional aspect of alcohol-related blackout. We focused here instead on the resilience of motor function to suppression by alcohol during the blackout state. We identified a sex-specific EEG marker associated with blackout history, and then found that the same marker was related to the frequency of episodes of certain non-REM parasomnias in both sexes. These findings suggest that these pathological states may share underlying dysfunction of motor inhibition, allowing for coordinated motor activity to persist during intoxication or sleep.

## Introduction

Alcohol-related blackouts (ARBs) are periods of acute intoxication during which a person maintains consciousness and motor control, but experiences total or partial amnesia of events (1–3). History of alcohol-related blackout is associated with an increased risk of injury resulting from alcohol use (4), premature death (5), and development of an alcohol use disorder (AUD) (6). Of those who drink, the literature indicates that somewhere between 37 and 75% of people have experienced a blackout in their lifetime (7–10). The literature also indicates that blackout is hereditary, even when controlling for drinking intensity (11, 12). Moreover, Wetherill and colleagues demonstrated that increases in activity in the bilateral frontal cortices and cerebellum, as measured with functional magnetic resonance imaging during an inhibitory task, were associated with future blackout in substance-naïve youth (13). Although the precise mechanisms of blackout are not fully understood, antagonism of NMDA receptors in the hippocampus has been proposed as a likely path of long-term potentiation (LTP) suppression (14–16).

Sleepwalking, which is also hereditary (17–19), shares behavioral features with blackout, including amnesia, behavioral disinhibition, and the persistence of motor function during a state of widespread neural suppression. Sleepwalking is categorized as one of three disorders of arousal (DoA), which are parasomnias defined by partial arousal during non-REM sleep (20). Data from our lab have linked DoA history to blackout likelihood, even when controlling for binge drinking, in a sample of adults who reported recent alcohol use (in preparation). Interestingly, this relationship appears restricted to females. Given the common behavioral features in blackout and DoA and the preliminary association between DoA and blackout history in women, we sought novel evidence for potential shared pathophysiology between DoA and blackout susceptibility. The aim of the current project was to identify neurophysiological characteristics associated with blackout history (Experiment 1) and history of DoA (Experiment 2), in an effort to determine whether there exist phenotypic risk indices shared between DoA and blackout susceptibility.

The human EEG spectrum can be decomposed into aperiodic and periodic signals (21). Aperiodic contributions to the signal are thought to contain information about excitatory/inhibitory (E/I) balance in the cortex. The aperiodic slope, which is regionally specific and modulated by pharmacological manipulation of global excitation or inhibition (22), has been used as a metric of cortical E/I balance (22–24). Slope has previously been shown to correlate with magnetic resonance spectroscopy measures of glutamate/GABA balance and with glutamate level in the prefrontal cortex, but not with GABA levels (25). When this aperiodic component is subtracted from the spectrum, the remaining peaks are thought to represent periodic (or oscillatory) synchronized patterns of neuronal firing in the cortex (21). Typically, the most prominent of these peaks is centered within a frequency range designated as alpha (7 – 13 Hz). Periodic activity in the alpha range reflects thalamo-cortical inhibition of task-irrelevant activity, and the location of the peak depends on the modality associated with the task at hand (26, 27). The most well-documented of these peaks is occipital alpha peak, detected at the highest amplitude from occipital electrodes, which inhibits task-irrelevant activity in the visual cortex (28). The amplitude of this peak increases when the eyes close, reflecting greater suppression of occipital activity when visual input is limited (29). In addition to the occipital alpha peak, the existence of a central sensorimotor alpha rhythm (mu-alpha) has been documented (30). The mu-alpha rhythm can be distinguished from the occipital alpha rhythm, indicating that they represent functionally distinct mechanisms (31). The mu-alpha rhythm can be modulated by the completion of a motor task or presentation of motor imagery, but not by eyes closing or opening (31). This particular pattern of modulation suggests that it serves a specific purpose inhibiting task-irrelevant activity in the motor cortex (31–34).

In the current paper, we analyzed resting-state EEG data from two samples collected at the University of North Carolina (UNC) at Chapel Hill to investigate the relationships between history of alcohol-related blackout, DoA, and characteristics of the resting-state EEG spectrum. We hypothesized that, after controlling for level of alcohol use, individuals with a history of blackout would exhibit differences in resting state EEG spectra compared to individuals with no such history, and that those differences would also be present in individuals with a history of DoA recruited as a separate sample. We predicted that in Experiment 1, individuals with a history of blackout would exhibit shallower slopes over the central primary motor cortex and reductions in mu-alpha power compared to controls with no history of blackout, indicating reduced inhibition in the region and a local shift in E/I balance toward excitation. In Experiment 2, we predicted that these same measures would be correlated with DoA severity such that shallower slopes and lower mu-alpha power would be associated with more severe DoA. Overall, we hypothesized that both alcohol-related blackout and DoA would be associated with a relative shift from inhibition towards excitation in the primary motor cortex at rest.

## Materials and Methods

The current project comprises two studies conducted at UNC Chapel Hill. The first was a multi-session project involving the administration of non-invasive neurostimulation (Experiment 1), and the second was a single-session study involving only the collection of resting-state EEG data and self-report questionnaire data (Experiment 2). All research activities were approved by the UNC Office of Research Ethics prior to the start of data collection.

### Experiment 1 (ARB)

#### Participants

Participants were recruited from the Chapel Hill area and surrounding cities for a multi-session behavioral, EEG, and neurostimulation study. The analyses described here included only data collected during the first session, before any neurostimulation was administered. Participants were recruited using online advertisements and flyers, and interested individuals were eligible if they were between the ages of 22 and 40, were native English speakers, were medically healthy, and held a high-school diploma or equivalent. Exclusion criteria included any contraindications to non-invasive neurostimulation, lifetime history of a substance use disorder, neurological or psychiatric disease, color blindness or related impairments, or past-month psychoactive drug use. Of those included in the following analyses, 50.9% were female and mean age was 28.11 (± 5.13).

#### Resting-state EEG data collection

EEG data were collected at a sampling rate of 1000 Hz using high-definition 128-channel EGI hydrocel geodesic nets (Magstim EGI, Eugene, OR). For each participant, head circumference, nasion-inion distance, and pre-auricular-pre-auricular distance were measured to determine net size and vertex location for net placement. Nets were soaked in a solution of KCl and water for 5 minutes to increase electrode conductivity, then placed on the participant by a trained researcher as recommended by the manufacturer. Electrode voltages were amplified using a Net Amps 400 amplifier and recorded using Net Station Acquisition software (version 5.4). Electrode impendences were checked prior to the beginning of data collection, and were minimized with the administration of additional KCl solution locally when necessary. Participants were situated in an electrically-shielded chamber during the duration of data collection to minimize the influence of outside electrical noise.

The resting-state data collection period was split into “eyes open” and “eyes closed” sections, each 7 minutes long. During the “eyes open” section, participants were instructed to look at a fixation cross on the screen in front of them. During the “eyes closed” section, they were instructed to close their eyes for the duration of the 7-minute block. Prior to the data collection period, participants were instructed to keep their heads still, minimize tension in the facial muscles, and avoid movement of the facial muscles and eyes.

#### Resting-state EEG data pre-processing

EEG datasets were individually pre-processed after collection using Net Station Tools software (version 5.4). For each dataset, 0.1 Hz high-pass and 100 Hz low-pass filters were applied. Datasets were then segmented into the 7-minute “eyes open” and 7-minute “eyes closed” epochs. Channels with excessive noise or poor scalp connection were visually identified by a trained researcher and marked for rejection. Baseline correction was applied using a baseline period of 1000 ms prior to segment start, unless the intended baseline period was unsuitable (e.g., included an eye blink) in which case a 1000 ms nearby period was chosen. Channels marked for rejection were then interpolated from surrounding channels, and the data were re-referenced from the vertex to a global average reference. Datasets were exported for cleaning and processing in MATLAB.

Datasets were cleaned using EEGLAB for MATLAB (35) as described by Robertson et al. (23) and reiterated here. Electrode locations were converted to the 10-10 international system (36), reducing the number of channels used in analysis from 128 to 71. This conversion did not involve averaging channels, although scalp topographies may have been affected by the interpolation used to replace rejected channels during preprocessing. An independent component analysis (ICA) was run on each epoch and artifacts were marked using the MARA plugin for EEGLAB (37). A trained researcher verified MARA’s categorizations for components accounting for more than 1% of the epoch variance, and modified when needed. Components that were deemed artifactual were removed.

#### Resting-state EEG data analysis

Cleaned EEG datasets were analyzed using built-in MATLAB functions, the Fieldtrip toolbox for MATLAB (38), the Signal Processing toolbox for MATLAB, and the fitting oscillations & 1/f (FOOOF) toolbox for python using a MATLAB wrapper (21). Scripts used to call these toolboxes for analysis were written by the research team. “Eyes open” and “eyes closed” epochs were analyzed separately. For each epoch, data were bandstop filtered from 58 to 62 Hz to minimize 60 Hz line noise (39). Power spectral densities (PSD) for each electrode of interest (Cz, C3, C4) were calculated across the epoch using Welch’s method (pwelch function). These electrodes were chosen because they are positioned over or near the primary motor cortex, with C3 and C4 generally corresponding to the right and left hands, respectively (40, 41). Epochs were divided into 1000ms segments with 50% overlap and analyzed using a Hamming window (23). Welch PSD outputs for each channel of interest were used as the inputs for the FOOOF model.

The model was run using custom MATLAB scripts, and “eyes closed” and “eyes open” datasets were run separately for each channel. The FOOOF models were run with the following parameters: maximum number of peaks 4, frequency range 2-50 Hz, peak threshold 2.0, minimum peak height 0.0, peak width limits 1-7 Hz, and aperiodic mode fixed. For each channel and condition (eyes open vs. closed), slope, individual peak alpha frequency, peak alpha power, and peak alpha bandwidth were output.

#### Variables

A series of questionnaires were administered during this experiment. The questionnaires relevant to this project were the Carolina Alcohol Use Pattern Questionnaire (CAUPQ) (42) and the Alcohol-Related Blackout Questionnaire (ARBQ). The CAUPQ assesses alcohol use under the age of 18, between ages 18 and 21, and during the past year. To approximate number of binges before the age of 18, we transformed the CAUPQ item assessing binges under 18 from an ordinal to a continuous scale as described by Elton et al. (42). To quantify current drinking patterns, CAUPQ responses were used to calculate “binge score.” (43). Binge score includes typical drinking rate, number of episodes of drunkenness during the past 6 months, and percentage of drinking days approached with the intention of getting drunk.

The ARBQ captures number of partial and total blackouts in the lifetime, before age 18, between the ages of 18 and 21, and during the past year. For both partial and total blackouts, participants are asked whether they’ve ever experienced that type of episode (“yes” or “no”), and if “yes” they are asked how many times during each period of interest. While this questionnaire has not been validated yet, Cronbach’s alpha in the sample was 0.958. For the primary analyses performed here, participants were categorized according to binary blackout history. Individuals who endorsed no lifetime history of partial or total blackouts were categorized as “no blackout,” individuals who endorsed any lifetime history of partial blackouts and no lifetime history of total blackouts were categorized as “partial blackout,” and individuals who endorsed any lifetime history of total blackouts (regardless of partial blackout history) were categorized as “total ± partial blackout.” In addition to the items directly referring to total and partial blackouts, the ARBQ also includes 3 items assessing specific features of the blackout experience. These include becoming physically lost while drinking, being unable to remember something one did while drinking, and recalling forgotten memories from when one was drinking. Responses corresponding to all 5 items regarding blackout episodes across the lifetime were averaged to create a continuous lifetime blackout score.

#### Statistical analyses

All statistical analyses were performed in SPSS version 29.0.1.0. A series of MANCOVAs were run to assess whether there was any relationship between blackout history, sex, and resting-state EEG characteristics. Each model included sex and blackout history (no, partial only, total ± partial) as factors and past year binge score (43), number of binges before age 18, and age as covariates. For each channel of interest, peak alpha power and peak alpha bandwidth were taken as dependent variables. Three MANCOVA models were tested to assess characteristics of the alpha peak at the target channels, with a threshold for significance at p ≤ .05 (two-sided). Additionally, because slope values at each electrode were strongly correlated with one another but not correlated with peak alpha power, a MANCOVA assessing aperiodic slope at C3, C4, and Cz was planned using the same factors and covariates. As described in the Results section, one individual was excluded from this analysis for poor FOOOF aperiodic model fit, which brought the smallest group size down to three. As this was equal to the number of dependent variables in the model, we chose to use a multivariate multiple regression model with composite lifetime blackout score as an independent variable instead. Regressions were run separately in males and females, and independent variables included were past year binge score, number of binges before age 18, age, and lifetime blackout score. Dependent variables were aperiodic slope at C3, Cz, and C4. In total, three MANCOVA models and two multivariate multiple regression models were tested. For all MANCOVA models, Box’s M and Levene’s tests were used to validate the assumptions of equal covariance matrices and homoscedasticity. For regression models, variance inflation factors were used to test for violations of multicollinearity and residuals were plotted for each dependent variable to test for violations of normality.

Custom SPSS syntax was used to test the assumption of homogeneity of regression slopes for each covariate. Violations were not found unless noted in the Results section. We specifically chose to use data collected during the “eyes open” resting-state condition for these analyses, as we suspected that volume conduction of the larger occipital alpha peak would obscure any group differences in mu-alpha power in the “eyes closed” condition (28).

### Experiment 2 (DoA)

#### Participants

Participants were undergraduate students enrolled in an introduction-level Psychology course at UNC Chapel Hill, recruited using the UNC Sona participant pool. All participants were age 18 or older. A sample of 40 was recruited, with 20 individuals recruited for any history of DoA episodes and 20 individuals recruited as controls. An issue with the collection of demographic information from this sample precluded analyses including age, sex, or ethnicity. The majority of participants were between age 18 and 22, and more than half were female or female-presenting.

#### Resting-state EEG data collection and analysis

Resting-state EEG data were collected using the same protocol used in Experiment 1, with minor differences in data processing. Specifically, during preprocessing with Net Station, rejected channels were interpolated before baseline correction. This adjustment was made to account for changes in dataset quality between Experiments 1 and 2 associated with age of the EEG nets. Additionally, during cleaning a 1 Hz high-pass filter was applied via EEGLAB to improve the ICA results (44). Data analysis was performed exactly as described for Experiment 1.

#### Variables

The Munich Parasomnia Screening was administered in this sample to assess DoA history and frequency (45). For each parasomnia, respondents indicate whether they have ever experienced that behavior, whether they currently experience it, and whether episodes were self-observed or observed by others. If they report that they formerly experienced that parasomnia, they are asked how many years have passed since the last occurrence. If they report that they currently experience that parasomnia, they are asked how frequently (1: very seldom – less than once a year, 2: seldom – once or several times a year, 3: sometimes – once or several times a month, 4: frequently – once or several times a week, or 5: very frequently – every or almost every night). For participants who reported currently experiencing episodes of sleepwalking, confusional arousals, and/or sleep terrors, we used the frequency of the most predominant episode type as the variable of interest. For example, a response of two for sleepwalking and four for confusional arousals would result in a DoA frequency score of four.

#### Statistical analyses

All statistical analyses were performed in SPSS version 29.0.1.0. As these individuals were recruited for history of DoA and without regard to drinking history, this dataset was underpowered for the blackout analyses performed in Experiment 1. Very few individuals in this sample reported any history of binge drinking or blackouts of either type. For this reason, bivariate correlations were used to identify whether any relationship existed between central peak alpha power, central aperiodic slope, and frequency of DoA episodes. Because the DoA frequency variable used here is ordinal, spearman’s ρ values are reported (46). Nonparametric correlations were tested with a threshold for significance at p ≤ .05 (two-sided).

## Results

### Experiment 1 (ARB)

Of the N = 67 participants from whom data was collected, N = 55 were included in the analyses described here. Eight were excluded for missing data relevant to the planned analyses. Three were excluded for reporting no lifetime history of alcohol consumption. Finally, one individual was excluded from the analyses for reporting that they typically consumed over 90 drinks per drinking day, which we deemed extremely unlikely to be accurate. One individual was excluded from the periodic analyses for failure of the FOOOF model to detect a peak within 7 and 13 Hz at any of the target electrodes. In the periodic analyses, 53 individuals were included in the analyses using channels C3 and Cz, and 52 included in the analysis using channel C4. One individual exhibited a poor model fit at C4 with all parameters tested (R^2^ = 0.430, error = 0.0892 for final model). Upon visual inspection, this was due to high frequency activity influencing the aperiodic component. The alpha peak was fit well compared to the original spectrum, so this individual was included in analyses concerning the alpha peak characteristics, but excluded from the slope analysis (N = 54 for slope analyses, 27 males and 27 females). Means and frequencies for relevant variables are available in **Table 1** for the 55 participants in this sample.

**Table 1:** Mean (±SD) values for relevant variables in Experiment 1.

| *Variable* | *N (%) or Mean (±SD)* | *Female* | *Male* |
| --- | --- | --- | --- |
| Age | 27.93 (±5.10) | 27.43 (±5.35) | 28.44 (±4.87) |
| Female | 28 (50.9) | - | - |
| Binge score | 13.89 (±25.21) | 9.59 (±10.22) | 18.36 (±34.22) |
| Adolescent binges | 14.55 (±44.16) | 16.91 (±52.48) | 12.09 (±34.30) |
| Lifetime blackout score | 2.37 (±2.07) | 2.45 (±1.81) | 2.28 (±2.34) |
| Any history of partial blackout | 41 (74.5) | 24 (85.7) | 17 (63.0) |
| Any history of total blackout | 22 (40.0) | 10 (35.7) | 12 (44.4) |
| Any history of blackout | 42 (76.4) | 24 (85.7) | 18 (66.7) |
| Days since last drink | 92.78 (±431.68) | 10.92 (±12.09) | 171.36 (±599.52) |

The following are the fit indices for the datasets included in the periodic analyses for each target electrode, according to the FOOOF outputs. The mean *R*^2^ for the included spectra at Cz was 0.996 (minimum 0.984, maximum 0.999) with a mean error value of 0.0350 (min 0.0121, max 0.0992). For C3, the mean *R*^2^ for the included spectra was 0.992 (min 0.927, max 0.999) with a mean error value of 0.0379 (min 0.0141, max 0.107). For C4, the mean *R*^2^ for the included spectra was 0.981 (min 0.430, max 0.999) with a mean error value of 0.0386 (min 0.0142, max 0.112). Excluding the aforementioned participant for the slope analysis, the mean R^2^ for the included spectra at C4 was 0.992 (min 0.878, max 0.999) with a mean error value of 0.0374 (min 0.0142, max 0.112). The following are the fit indices for the datasets included in the aperiodic analyses, according to the FOOOF outputs. The mean R^2^ for the included spectra at Cz was 0.997 (minimum 0.984, maximum 0.999) with a mean error value of 0.0351 (min 0.0121, max 0.0992). For C3, the mean R^2^ for the included spectra was 0.993 (min 0.927, max 0.999) with a mean error value of 0.0379 (min 0.0141, max 0.107). These R^2^ and mean error values validate that the FOOOF model parameters used allowed for good model fits of the raw spectra at the target electrodes.

### Peak alpha power and peak alpha bandwidth by blackout group and sex

First, we sought to determine whether the oscillatory activity represented by the alpha peak differed according to group at each electrode. Peak power and bandwidth account for the magnitude of that activity, so for each electrode we performed a multivariate analysis to determine whether blackout predicted either periodic metric. As an overview, we found that history of blackout was associated with reductions in alpha peak power in females only, and this relationship was strongest centrally (Cz). No relationship was found between alpha peak bandwidth and blackout history. Results for alpha peak parameters at each electrode are described in the following sections.

#### C3

The MANCOVA model including peak alpha power and peak alpha bandwidth at C3 indicated that there was a trending interaction between sex and blackout history (Pillai’s trace = 0.183, *F*_(4, 88)_ = 2.22, *p* = .073) in predicting the dependent variables. Thus, we did not detect significant differences between groups in alpha peak characteristics at C3. Levene’s tests indicated that the assumption of homoskedasticity was not violated for bandwidth, but it should be noted that it was violated for power (power: *F*_(5, 47)_ = 2.60, *p* = .037; bandwidth: *F*_(5, 47)_ = 0.26, *p* = .93). In a follow-up ANOVA with peak bandwidth as at C3 the dependent variable, the omnibus model was not significant (*F*_(8, 44)_ = 1.64, *p* = .14) and sex was a significant predictor (*F*_(1, 44)_ = 4.94, *p* = .031), such that peak bandwidth was lower in females than in males. In a follow-up ANOVA with peak power at C3 as the dependent variable, the omnibus model was not significant (*F*_(8, 44)_ = 1.82, *p* = .10) and the interaction between sex and blackout history was a significant predictor (*F*_(2, 44)_ = 4.26, *p* = .020). In females, peak alpha power at C3 was reduced in the total ± partial blackout group (p = .019) and in the partial blackout group (p = .032) compared to the no blackout group, while there were no discernible differences between groups in males. Mean alpha peak power values at C3 are displayed by group and sex in **Figure 1A** and mean peak bandwidths are displayed according to sex in **Figure 2A**.

**Figure 1:**
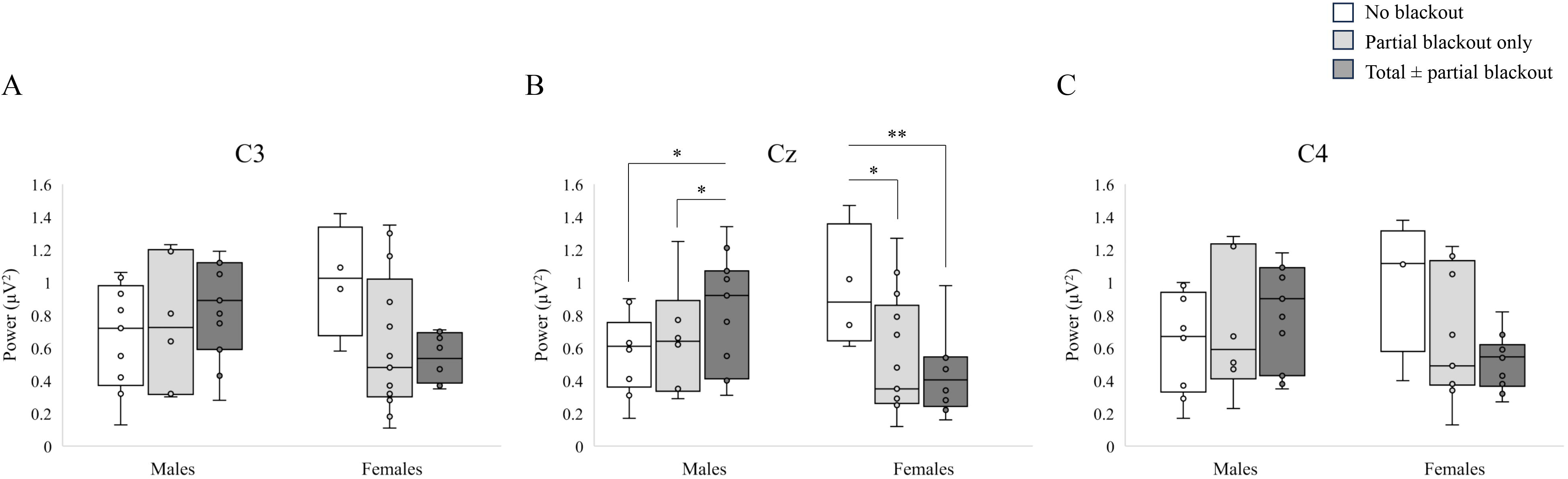
Peak alpha power at each target electrode plotted according to sex and blackout group (Experiment 1). Results of pairwise comparisons of simple main effects between groups following univariate ANCOVAs with significant omnibus results displayed. * p < .05. ** p < .01. A: Peak alpha power at C3 (left primary motor cortex). N = 53. B: Peak alpha power at Cz (central primary motor cortex). N = 53. C: Peak alpha power at C4 (right primary motor cortex). N = 52.

**Figure 2:**
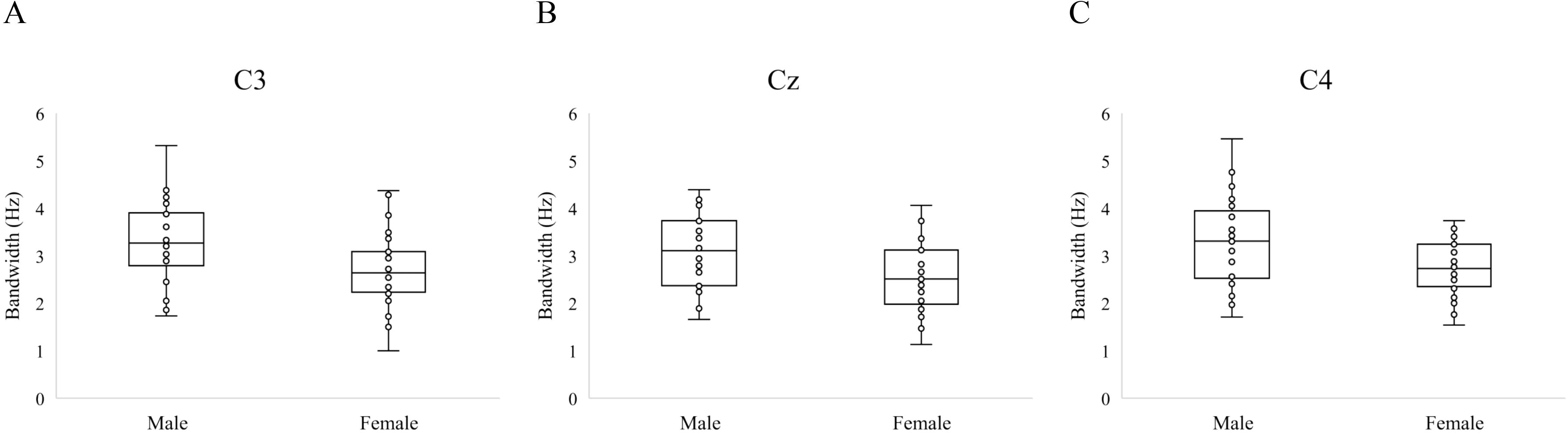
Alpha peak bandwidth at each target electrode plotted according to sex (Experiment 1). Alpha peak bandwidth at C3 (left primary motor cortex). N =53. B: Alpha peak bandwidth at Cz (central primary motor cortex). N = 53. C: Alpha peak bandwidth at C4 (right primary motor cortex). N = 52.

#### Cz

The MANCOVA model including peak alpha power and peak alpha bandwidth at Cz indicated that there was a significant interaction between sex and blackout history (Pillai’s trace = 0.276, *F*_(4, 88)_ = 3.52, *p* = .010) in predicting the dependent variables at Cz. In a follow-up ANOVA with peak bandwidth as the dependent variable, the omnibus model was not significant (*F*_(8, 44)_ = 1.87, *p* = .089). In a follow-up ANOVA with peak power at Cz as the dependent variable, the omnibus model was significant (*F*_(8, 44)_ = 3.12, *p* = .007) and the interaction between sex and blackout history was a significant predictor (*F*_(2, 44)_ = 7.56, *p* = .002). This interaction indicated females who reported experiencing a blackout had reduced peak alpha power at Cz compared to those who had never reported a blackout, and that this was not the case for males in this sample. This was true even when controlling for age, current binge score, and approximate number of binges before age 18, suggesting that this reduction in alpha power is specific to blackout rather than being reflective of heavy alcohol use. Mean alpha peak power values at Cz are displayed by group and sex in **Figure 1B** and mean peak bandwidths at Cz are displayed according to sex in **Figure 2B**. Univariate tests using the estimated marginal means of alpha peak power indicated that there were significant differences according to blackout group in males (*F*_(2, 44)_ = 3.56, *p* = .037), with the total ± partial blackout group having increased peak alpha power compared to the no blackout (p = .020) and partial blackout groups (p = .041). There were significant univariate differences among females (*F*_(2, 44)_ = 4.36, *p* = .019), with groups differing in the opposite direction. Pairwise comparisons indicated that females with no history of blackout differed significantly from females with a history of total ± partial blackouts (p = .006), and from females with a history of partial blackout only (p = .018). These groups differed such that alpha power was significantly lower in both blackout groups compared to controls, but the two blackout groups did not differ from one another.

#### C4

The MANCOVA model including peak alpha power and peak alpha bandwidth at C4 indicated that there was a significant main effect of sex (Pillai’s trace = 0.166, *F*_(2, 42)_ = 4.19, *p* = .022) and a significant interaction between sex and blackout history (Pillai’s trace = 0.207, *F*_(4, 86)_ = 2.48, *p* = .050) in predicting the dependent variables at C4. In a follow-up ANOVA with peak bandwidth as the dependent variable, the omnibus model was not significant (*F*_(8, 43)_ = 1.61, *p* = .15) and sex was a significant predictor (*F*_(1, 43)_ = 8.37, *p* = .006). In a follow-up ANOVA with peak power at C4 as the dependent variable, the omnibus model was not significant (*F*_(8, 43)_ = 1.83, *p* = .097) and the interaction between sex and blackout history was a significant predictor (*F*_(2, 43)_ = 4.89, *p* = .012). In pairwise comparisons, females with a history of total ± partial blackout had significantly lower peak alpha power compared to females with no history of blackout (p = .018). These results suggested that sex and blackout history may contribute to alpha peak characteristics at C4, but post-hoc comparisons did not meet our threshold for significance. Mean alpha peak power values at C4 are displayed by group and sex in **Figure 1C** and mean peak bandwidths at C4 are displayed according to sex in **Figure 2C**. The pattern displayed here was similar to that seen in electrodes C3 and Cz.

Of the three MANCOVA models tested, only the one including power and bandwidth at Cz provided sufficient statistical evidence of sex-specific group differences according to blackout history. To further explore this sex-dependent relationship between blackout history and central alpha peak power, we calculated bivariate correlations between lifetime composite blackout score (derived from the ARBQ; see Methods) and alpha peak power at Cz. These correlations were run separately in males (n = 26) and females (n = 27). In females, there was a significant correlation between lifetime composite blackout score and alpha power at Cz (ρ = −0.480, p = .011). This relationship was not significant in males (Cz: ρ = 0.230, p = .26). Peak alpha power at Cz is plotted against lifetime blackout score in **Figure 3**. This is consistent with the MANCOVA results described previously, as it indicates the existence of a relationship between blackout susceptibility and peak alpha power at Cz in females specifically. It also suggests that this phenotype may exist on a spectrum, with lower magnitude alpha peaks associated with greater history of blackout.

**Figure 3:**
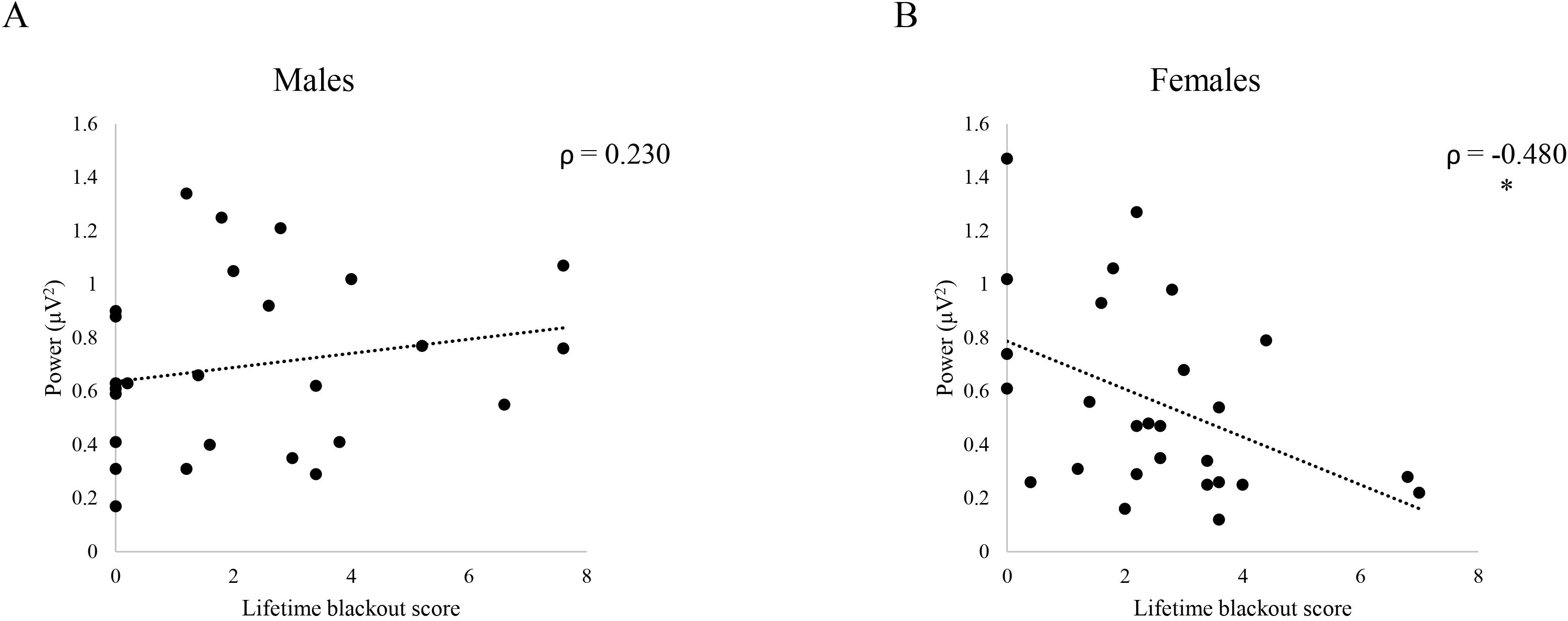
Peak alpha power at Cz (central primary motor cortex) plotted against lifetime blackout score (Experiment 1). Spearman’s ρ correlations reported. * p < .05. ** p < .01. *** p < .001 A: Relationship in males (n = 26). B: Relationship in females (n = 27).

With regard to the aperiodic slope analyses, the multivariate linear regression model run in males indicated that composite blackout score was the only significant multivariate predictor of slope at the target electrodes (Pillai’s trace = 0.473, *F*_(3,20)_ = 5.99, *p* = .004). ANOVAs testing model fit for each dependent variable indicated that fit was better than an intercept-only model for slope at C4 (*F*_(4, 22)_ = 3.45, *p* = .025) and C3 (*F*_(4, 22)_ = 2.88, *p* = .046), but not at Cz (*F*_(4, 22)_ = 1.58, *p* = .216). Unstandardized coefficients for each predictor are available for univariate and multivariate models in **Table 2**. Looking at specific predictors, lifetime blackout score was the only significant predictor of slope and only predicted it at C4 (t = −2.53, p = .019). The relationship between blackout score and slope at C4 was such that as blackout score increased, slope flattened, indicating a shift toward excitation (22). No individual variable was a significant predictor of slope at C3. The multivariate linear regression model run in females did not indicate that any of the independent variables included were significant multivariate predictors of slope at the target electrodes. Unstandardized coefficients for each predictor are available for univariate and multivariate models in **Table 3**. These results suggested that there exists a lateralized relationship between slope of the ipsilateral dominant primary motor cortex and blackout history in males specifically. This is in contrast with the periodic findings, which provided evidence for a relationship between blackout history and central peak alpha power in females. Together, these findings indicate that predisposition to blackout may be related to hyperexcitability of the motor cortex in both males and females, but that the nature of that disturbed E/I balance differs according to sex. In females, inhibition of the motor cortex, as operationalized as alpha power, is attenuated in those with a history of blackout, and in males, E/I balance is shifted toward excitation, as operationalized as aperiodic slope, in those with greater history of blackout. The relationships between blackout score and slope at each of the target electrodes are plotted for males in **Figure 4A** and for females in **Figure 4B**.

**Figure 4:**
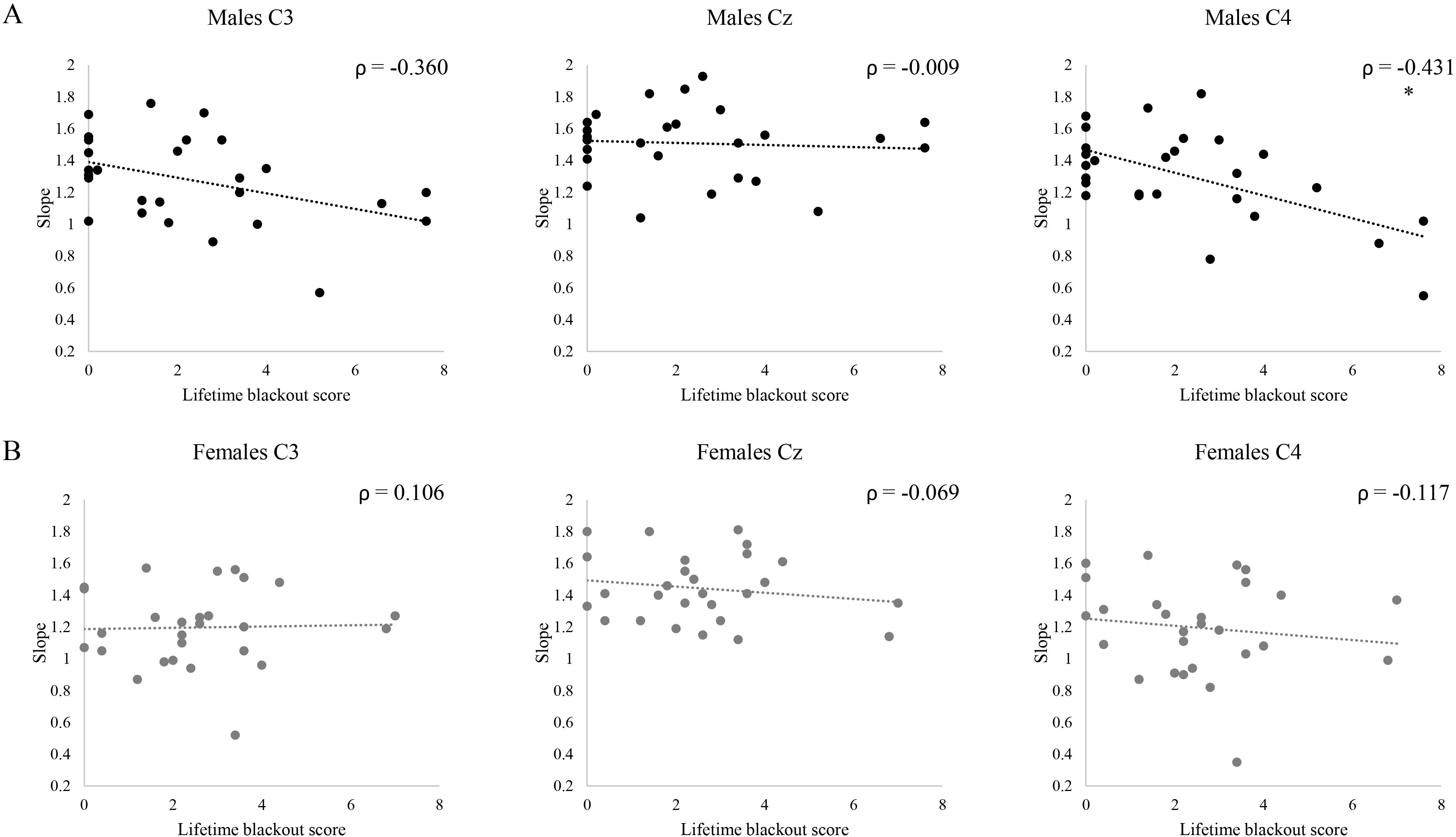
Aperiodic slopes at each electrode plotted against lifetime blackout score (Experiment 1). Spearman’s ρ correlations reported. * p < .05. ** p < .01. *** p < .001 A: Relationships in males (n = 27). B: Relationships in females (n = 27).

**Table 2:** Multivariate linear regression model results in males (n = 27; Experiment 1).

| *Predictor* | Univariate (C3) |  |  | Univariate (Cz) |  |  | Univariate (C4) |  |  | Multivariate |  |
| --- | --- | --- | --- | --- | --- | --- | --- | --- | --- | --- | --- |
|  | *B* | *t* | *Sig.* | *B* | *t* | *Sig.* | *B* | *t* | *Sig.* | *F*<br>(3, 20) | *Sig.* |
| Age | 0.00 | -0.11 | .99 | -0.003 | -0.37 | .72 | -0.004 | -0.35 | .73 | 0.11 | .95 |
| Binge score | -0.002 | -1.33 | .20 | -0.002 | -1.85 | .078 | -0.002 | -1.30 | .21 | 1.04 | .40 |
| Adolescent binges | -0.003 | -1.92 | .068 | -0.002 | -1.51 | .15 | 0.00 | 0.18 | .86 | 2.35 | .10 |
| **Lifetime blackout score** | -0.014 | -0.56 | .58 | 0.021 | 0.96 | .35 | **-0.065** | **-2.53** | **.019** | **5.99** | **.004** |

B: unstandardized regression coefficient

**Table 3:** Multivariate linear regression model results in females (n = 27; Experiment 1).

|  | Univariate (C3) |  |  | Univariate (Cz) |  |  | Univariate (C4) |  |  | Multivariate |  |
| --- | --- | --- | --- | --- | --- | --- | --- | --- | --- | --- | --- |
| *Predictor* | *B* | *t* | *Sig.* | *B* | *t* | *Sig.* | *B* | *t* | *Sig.* | *F*<br>(3, 20) | *Sig.* |
| Age | 0.00 | 0.038 | .97 | **-0.016** | **-2.08** | **.050** | -0.012 | -1.17 | .26 | 2.37 | .10 |
| Binge score | **-0.011** | **-2.24** | **.036** | -0.007 | -1.62 | .12 | **-0.016** | **-2.86** | **.009** | 2.70 | .073 |
| Adolescent binges | -0.001 | -0.75 | .46 | 0.00 | 0.35 | .73 | -0.002 | -1.71 | .10 | 2.97 | .057 |
| Lifetime blackout score | 0.018 | 0.67 | .51 | -0.012 | -0.53 | .60 | -0.001 | -0.047 | .96 | 0.64 | .60 |

B: unstandardized regression coefficient

To summarize the findings of Experiment 1, females with a history of blackout (partial or total) had reduced alpha power in the central primary motor cortex compared to those with no history, whereas blackout history was related to this marker in the opposite direction in males. This relationship in females was strongest at electrode Cz. The aperiodic analyses suggested that in males, slope at C4 is shifted toward excitation in those with greater propensity to experience blackout. Both of these markers are associated with disruptions in normative E/I balance, but it appears that deficient inhibition may be underlying this disruption in females, while it may reflect increased excitation in males.

#### Experiment 2 (DoA)

Of the N = 40 participants from whom data was collected, N = 36 were included in the analyses described here because four participants’ datasets were excluded for poor quality of EEG data. Of those 36, the FOOOF model failed to detect a peak between 7 and 13 Hz at channel C3 for one individual. As a result, 36 individuals were included in the analyses using channels Cz and C4, and 35 were included in the analyses using channel C3. Of the 36 participants included in these analyses, 21 reported any lifetime history of DoA. Of those 21, the median frequency of DoA episodes was 2.00 (seldom – once or several times a year).

According to the FOOOF output, the mean *R*^2^ for the included spectra at Cz was 0.996 (min 0.962, max 0.999) with a mean error value of 0.0305 (min 0.0140, max 0.0523). For C3, the mean *R*^2^ for the included spectra was 0.994 (min 0.927, max 0.999) with a mean error value of 0.0333 (min 0.0186, max 0.0585). For C4, the mean *R*^2^ for the included spectra was 0.992 (min 0.889, max 0.999) with a mean error value of 0.0365 (min 0.0150, max 0.0936). These R^2^ and mean error values validate that the FOOOF model parameters used allowed for good model fits of the raw spectra at the target electrodes.

In order to determine whether any relationships existed between DoA severity, alpha peak power in the motor cortex, and aperiodic slope in the motor cortex, we ran nonparametric bivariate correlations with the following variables: peak alpha power at Cz, peak alpha power at C3, peak alpha power at C4, slope at Cz, slope at C3, slope at C4, and recent DoA frequency. Spearman’s ρ values are available in **Table 4**. DoA frequency was significantly negatively correlated with alpha peak power at Cz (*p* = .004), C3 (*p* < .001), and C4 (*p* = .013). This means that more frequent parasomnia episodes were associated with lower central alpha peaks at all locations in this sample. DoA frequency is plotted against alpha peak power at each target electrode in **Figure 5**. Blackout history and alcohol use data were collected from this sample using the ARBQ and CAUPQ, respectively, but alcohol use was so low that we were underpowered to perform the alcohol-related analyses from Experiment 1 in this sample.

**Figure 5:**
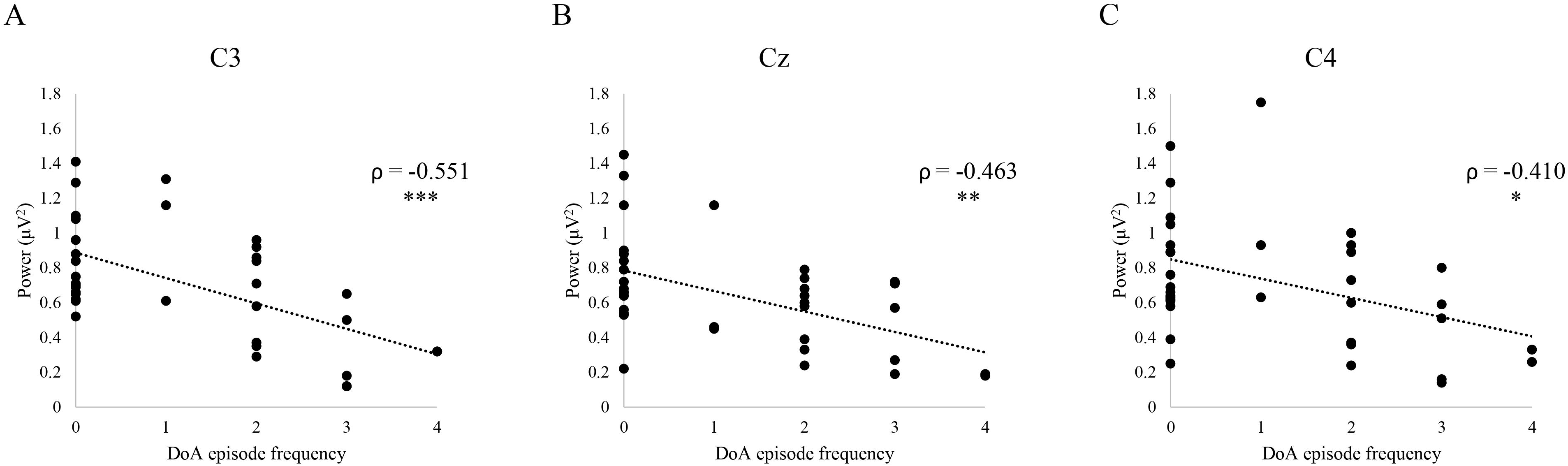
Peak alpha power at each target electrode plotted against recent frequency of disorder of arousal episodes (Experiment 2). Spearman’s ρ correlations reported. * p < .05. ** p < .01. *** p < .001 A: Peak alpha power at C3 (left primary motor cortex). N = 35. B: Peak alpha power at Cz (central primary motor cortex). N = 36. C: Peak alpha power at C4 (right primary motor cortex). N = 36. DoA: disorder of arousal.

**Table 4:** Spearman’s ρ values for the correlations between DoA episode frequency and measures related to central excitatory/inhibitory balance (Experiment 2)

|  | DoA frequency | Peak alpha power Cz | Peak alpha power C3 | Peak alpha power C4 | Slope Cz | Slope C3 |
| --- | --- | --- | --- | --- | --- | --- |
| DoA frequency | 1.000 |  |  |  |  |  |
| Peak alpha power Cz | **-0.463\*\*** | 1.000 |  |  |  |  |
| Peak alpha power C3 | **-0.551\*\*\*** | **0.627\*\*\*** | 1.000 |  |  |  |
| Peak alpha power C4 | **-0.410\*** | **0.582\*\*\*** | **0.738\*\*\*** | 1.000 |  |  |
| Slope Cz | -0.059 | 0.250 | 0.294 | **0.380\*** | 1.000 |  |
| Slope C3 | -0.118 | **0.345\*** | 0.276 | 0.281 | **0.650\*\*\*** | 1.000 |
| Slope C4 | 0.009 | 0.208 | 0.163 | 0.320 | **0.648\*\*\*** | **0.721\*\*\*** |

\* $p < .05$ ; \*\* $p < .01$ ; \*\*\* $p < .001$ ; DoA: disorder of arousal

## Discussion

When combined, the results of Experiments 1 and 2 supported our hypothesis that alcohol-related blackout and DoA share disturbances in motor E/I balance as a neurophysiological phenotype. In Experiment 1, we found that reductions in central alpha power in the motor cortex were associated with history of blackout in females, whereas in males, history of blackout was associated with a lateralized shift in aperiodic slope over the motor cortex. These measures both quantify cortical excitability, although they reflect different underlying constructs related to cellular inhibition and excitation (47). In Experiment 2, we found that central alpha power in the motor cortex was negatively correlated with frequency of DoA episodes. This is the same measure of inhibition found to be related to blackout history in females in Experiment 1, indicating that motor hyperexcitability may be a predisposing factor for both DoA and blackout.

### Central periodic alpha power and blackout

In Experiment 1, females with a history of any type of alcohol-related blackout had lower resting-state peak alpha power at central electrodes than did females with no blackout history, even when controlling for measures of alcohol consumption. Thus, this phenotype may be associated with risk of blackout in females. The alpha peak detectable over sensorimotor regions reflects the mu-alpha rhythm, and can be dissociated from the occipital alpha rhythm involved in inhibiting activity in the visual cortex (28, 31). Mu-alpha power is reduced during completion (or observation) of a motor task, and is related to excitability of the motor cortex as assessed with single-pulse TMS (30, 34, 48, 49). Within the framework of our hypothesis, differences in resting-state mu-alpha power may represent reduced inhibition of the motor cortex in females with a history of blackout. This reduced inhibition may be negligible at rest, but with sufficient intoxication may lead to relative sparing of motor function compared to other neural domains. This unbalanced suppression could allow for motor function to persist, as in the blackout state, while hippocampal and prefrontal activity are severely limited.

As this theoretical framing relies on a relationship between blackout history and motor <u>hyper</u>excitability, it seems paradoxical that males with a history of total blackout exhibited increased central peak alpha power on average (i.e., more inhibition) compared to the other groups. As discussed further below, aperiodic slope at C4 was negatively correlated with lifetime blackout score in males. It may be the case that in males, a severe shift toward excitation in the motor cortex is required to increase blackout susceptibility, and that this increase in alpha power is a compensatory mechanism in those on the most extreme end of the spectrum. This is supported by the fact that alpha power was increased only in the total ± partial blackout group, which you would expect to display the most severe E/I disturbance.

Aperiodic slope is a gross measure of E/I balance – it may be that as hyperexcitability becomes extreme enough to be detected by slope, thalamo-cortical and cortico-cortical inhibitory circuits are recruited to compensate. This supposition could be investigated in future work.

### Central aperiodic slope and blackout

In Experiment 1, aperiodic slope at C4 (right primary motor cortex) was negatively correlated with lifetime blackout score in males. This suggests that as lifetime exposure to blackout increases, slope flattens with unilateral specificity. Flattening of the slope across the scalp, including at central sites, has been associated with aging and with reductions in performance of cognitive tasks (50, 51). Slope is thought to represent E/I balance of the underlying cortex, with flatter slopes reflecting a shift toward excitation (22). This relationship between slope at C4 and blackout history indicates that as excitability of the motor cortex increases in males, propensity to blackout also increases. As described in relation to the central alpha peak in females, this hyperexcitability could predispose some individuals to blackout when the blood alcohol concentration (BAC) is elevated sufficiently. Alternatively, these data could indicate that exposure to the blackout state causes injury to the brain in males.

### Central periodic alpha power and DoA

In Experiment 2, we observed that resting-state peak alpha power at central electrodes, the same neurophysiological index which was associated with blackout history in females in Experiment 1, was negatively correlated with frequency of DoA episodes. This observation was consistent with previous findings in individuals with DoA and the similarity in associations across the two studies was consistent with our a priori hypotheses. Given the behavioral similarities between blackout and DoA episodes, we hypothesized that there may be neural mechanisms shared by the two states. These findings provide support for that notion, and indicate that reduced mu-alpha power at rest may be a shared predisposing factor.

Using single-pulse and paired-pulse TMS, Olivero et al. found evidence of impaired inhibition of the motor cortex at rest in sleepwalkers (52). Central alpha power is related to excitability of the motor cortex, so these findings are consistent with the extant literature (30, 49). As hypothesized with regard to blackout, we suspect that this reduced motor cortical inhibition may contribute to breakthrough motor function during non-REM sleep.

### Sex differences in blackout risk

The results of Experiment 1 indicated that some risk factors underlying alcohol-related blackout may be sex-dependent. The published literature on blackout is equivocal as to whether males or females are more prone to blackout episodes, or whether sex differences in risk beyond metabolic factors exist at all (7, 53–55). The results of this study indicate that insufficient periodic inhibition of the motor cortex may interact with the aforementioned metabolic factors to increase risk of blackout in some females.

Unlike the findings of Experiment 1, the relationship between central alpha peak power and DoA frequency was present in the sample as a whole in Experiment 2. Although sex differences in the behaviors that occur during parasomnia episodes have been documented, we found no evidence in the literature indicating that sex differences in mechanism or predisposing factors should be expected (56).

Perhaps the observation that this mechanism is sex-specific in blackout, but not parasomnia, reflects other sex-specific mechanisms having to do with alcohol exposure. For example, females are known to metabolize alcohol less efficiently than males do, and females have lower body weights and higher body fat percentages on average (57, 58). It could be that existing risk for blackouts, measured here as reduced alpha over motor cortex, interacts with this inefficient metabolization of alcohol to specifically increase risk for blackout in females.

### Limitations

We recognize that this project has several limitations. First, we interpreted these findings as being indicative of the existence of a predisposing phenotype that increases lifetime blackout risk. This was informed by the literature suggesting that blackout risk is heritable and associated with aberrant neurophysiology prior to initiation of substance use (11–13). However, it must be noted that assigning directionality to these effects is beyond the scope of these data. A longitudinal study would be required to discern whether reduced mu-alpha power is a risk factor, or a consequence of alcohol-related blackout(s). Regardless of the directionality of this relationship, though, these findings are novel and provide support for future work.

Additionally, the sample sizes for Experiments 1 and 2 were relatively small, and our groups for the analyses used in Experiment 1 were unbalanced. This sample had a particularly high prevalence of lifetime blackout (75.9%), reducing the size of our non-blackout groups. Ironically, the sample collected in Experiment 2 had a particularly low prevalence of lifetime blackout (25%). These concerns, however, are somewhat mitigated by a power analysis, which indicated that the analyses completed for Experiment 1 were sufficiently powered, and by Box’s test, which indicated that the assumption of equivalence of covariance matrices was not violated for any of the MANCOVA models.

## Conclusions

The results of this project supported our hypothesis that alcohol-related blackout and DoA share neurophysiological risk factors. Our findings indicate that one of those shared factors may be reduced central peak alpha power at rest. This neurophysiological index, indicative of increased excitation of the motor cortex, was correlated with frequency of DoA episodes in the sample as a whole in Experiment 2, and with history of blackout specifically for females in Experiment 1. Interestingly, for males, history of blackout was associated with a different index of increased excitation over the motor cortex; flatter aperiodic slope. These findings suggest that future research should explore neurophysiological phenotypes relating to excitability of the motor cortex in participants with both parasomnias and blackouts related to alcohol, ideally in longitudinal designs which can establish this risk phenotype as an antecedent to alcohol-related blackout.

## Data Availability

The corresponding author can make data available upon request.

## Acknowledgments

Present address of MMR: Department of Psychiatry & Behavioral Sciences, Duke University, Durham, NC, USA.

## Disclaimers

None.

## Author Contributions

Data analysis and manuscript writing were done by GME. Data for experiment 1 were collected by MMR, and data for experiment 2 were collected by GME and CEL. EEG pre-processing for experiment 1 was done by MMR, and by GME for experiment 2. This project was designed by GME and CAB with assistance from DLR and MAS. All authors read and edited this manuscript.

